# Systematic review of COVID-19 autopsies: accelerated hyaluronan concentration as the primary driver of morbidity and mortality in high-risk COVID-19 patients: with therapeutic introduction of an oral hyaluronan inhibitor in the prevention of “Induced Hyaluronan Storm” Syndrome

**DOI:** 10.1101/2020.04.19.20071647

**Authors:** Michael A. Mong, Jacob A. Awkal, Paul E. Marik

## Abstract

To date, the fundamental drivers of the morbidity and mortality in COVID-19 remain uncertain. Clinicians worldwide appear to be at a loss to know how to prevent and treat the severe respiratory distress in these patients effectively. Consequently, the fundamental mechanisms leading to death in high-risk patients need to be discovered and addressed with urgency. The post-mortem autopsy remains an essential part of both discovering the cause of death in a particular individual, but also in advancing the science and treatment of disease, especially in the case of novel pathogens such as SARS-CoV-2^[2]^. The goal of an autopsy is to discover the cause of death (COD) using a macro/microscopic investigation. Because lung weight is often affected by the cause of death and the last breath occurs very near if not at death, the evaluation of the lungs is one of the starting points of any COD investigation^[3]^. A comprehensive search was performed to systematically review all reported autopsy findings in COVID-19 patients with respect to lung weights and histologic findings. We then compared these findings with the results of a targeted literature review of hyaluronan in relationship to acute respiratory distress syndrome (ARDS). In total, data from 38 autopsies were identified. From this group, 36 autopsies of COVID-19 patients were selected for detailed review and statistical analysis. The average lung weight of those who were determined to have died as a result of SARS-CoV-2 was 1683g approximately 3.2 times the normal lung weight. Hyaline membranes were consistently identified on histologic sections. A review of the literature reveals that markedly elevated lung weights and hyaline membranes and have been associated with the pathophysiology of ARDS since 1967. However, the key role key of hyaluronan in driving the morbidity and mortality of the condition has heretofore not been fully recognized. We propose that the induced hyaluronan storm syndrome or IHS, is a model that addresses the heretofore perplexing respiratory failure that is the proximal cause of death. An aggressive research effort should be undertaken to discover why the majority of individuals who are exposed to the virus are minimally symptomatic, while a minority of high-risk individuals rapidly progress to respiratory failure and death.

*“You may take notes for 20 years, from morning to night at the bedside of the sick, upon the diseases of the viscera, and all will be to you only a confusion of symptoms*…*a train of incoherent phenomena. Open a few bodies and this obscurity will disappear*.*” - Marie-François-Xavier Bichat (1771–1802), “The Father of Histology”*^*[1]*^.

## Introduction

To date no coherent model has emerged to explain, prevent, or treat the rapid respiratory failure that is the proximal cause of death in a minority of 2019 novel coronavirus (2019-nCOV; SARS-CoV-2; COVID-19) patients. Similarly, this remains true for the acute respiratory distress syndrome (ARDS) first described by Ashbaugh in 1967^[4]^ seen in the severe acute respiratory syndrome (SARS) outbreak in 2002, and the Middle East respiratory syndrome (MERS) in 2012. Clearly, a small subset of infected individuals rapidly progress, often in a few hours to a few days, from initial symptoms of fever and dyspnea, to full-blown respiratory failure requiring mechanical ventilation, while others remain essentially asymptomatic. Once placed on a ventilator, the death rate has been reported to be between 37 to 62 percent and up to 80 percent depending on age and other risk factors^[5]^. Currently, the explanation offered to explain this rapid chain of events has been the cytokine storm syndrome (CSS)^[6,7]^. However, this model is lacking in some respects, and does not readily lead to a broad based hypothesis and treatment protocol that can be tested immediately. Using a systems biology framework^[8]^, we conducted a comprehensive review of autopsy reports relating to SARS-CoV-2 deaths, and reviewed the literature concerning hyaluronan with respect to acute respiratory distress syndrome (ARDS). We found that there is a consistent and statistically significant increase in the reported lung weights at autopsy in confirmed SARS-CoV-2 victims. Most significantly, in the first report of two complete autopsies in SARS-CoV-2 in the English language literature, the observed lung weight of a victim with a positive nasopharyngeal and bilateral lung parenchymal swab, by rRT-PCR for SARS-CoV-2, weighed 2,452g - approximately 4.6 times the normal adult lung weight. These lungs were described at autopsy as a “bag of water” (personal communication)^[9]^. In sharp contrast, and most importantly, a 42-year-old obese man was determined to have died with SARS-CoV-2 but not because of SARS-CoV-2. At postmortem examination, the nasopharyngeal swab tested positive for SARS-CoV-2 but there was a negative lung parenchymal swab, with normal lung weights and findings of acute bronchopneumonia.

Modig and Hällgren in 1989, elegantly demonstrated that hyaluronic acid (HA) can be induced within minutes in alveolar fluids and leads to a dose dependent reduction in the pulmonary oxygenation index, PaO_2_/FaO_2_. They further demonstrated up to an 82 times increase in alveolar HA concentration in ARDS patients compared to controls. Stating that “hyaluronic acid is the most powerful water(edema)-binding substance in the body”, they put forth a “humoral” and “biochemical-physiological” hypothesis of ARDS^[10]^. Recently, others have detailed the central role of HA in human respiratory disease, in which viral infections lead to marked and rapid increase in HA concentrations^[11]-[13]^. This may serve to confirm the unique cause of death in the postmortem examination, aforementioned by Barton et al., whereby the super-normal outpouring of HA over a short time interval within the confined space of the human chest cavity leads to acute respiratory distress. We propose to define a new model of the induced hyaluronan storm (IHS) syndrome, as a comprehensive, specific, and actionable paradigm to frame and address the global crisis and specifically, to prevent death in COVID-19 high-risk groups.

In 2013, McKallip, et al. reported an *in vitro* and *in vivo* model of ARDS stating that “targeting hyaluronic acid synthesis might be a novel target for the treatment or reduction of induced lung injury”. Using an elegant combination of cell based and murine studies, they demonstrated that it was possible to profoundly suppress the production of HA by inhibiting the three isoenzymes of HA synthase (HAS-1, HAS-2, and HAS-3) with 4-methylumbelliferone (4-MU)^[14]^.

A follow up study by the same group went on to show that 4-MU reduces the expression of HAS-1, HAS-2, and HAS-3, and reduced levels of HA in the lungs of Staphylococcal enterotoxin B (SEB)-exposed mice and that 4-MU treatment yields a reduction in SEB-induced lung permeability and reduced cytokine production^[15]^.

Bray et al. in 1991 demonstrated in a bleomycin-induced lung injury model, maximal HA content was reached after seven days and was 14.6 times the normal value^[16]^. More recently, Reeves et al. in 2020, showed that human lung fibroblasts (HLFs) treated with viral mimetic polyinosine-polycytidylic acid, a Toll-like receptor 3 agonist, encourages the accumulation of HA-rich extracellular matrix (ECM) and enhances monocyte and lymphocyte binding. It was shown that the activity of mast cells (innate immune cells), particularly in acute respiratory infections such as RSV, can lead to the formation of a HA-enriched ECM. This further emphasizes that RSV infections of HLFs encourage inflammation via HA-dependent mechanisms that enhance mast cell protease expression through direct contact with the ECM^[17]^. Furthermore, HA and its various degradation products *in vitro* have been shown to be a significant contributor to the immunomodulatory functions of the ECM in both acute and chronic respiratory diseases.

In 2015, Paul Bollyky, M.D., PhD. and Nadine Nagy PhD. et al., researchers at Stanford, extensively reviewed the use of 4-MU in both animal and human studies. This was accomplished by investigating its mechanism of action, pharmacokinetics, and safety, with respect to cancer and autoimmunity. These researchers reported eight human studies dating back to 1978 involving HA and cancer. They concluded that 4-MU has the potential of becoming a long-term adjunctive therapy for a myriad of indications^[18]^.

## Methods

### Study Type

A systematic comprehensive review of COVID-19 autopsy reports and scientific literature using key search terms.

### Inclusion Criteria

The inclusion criteria of this study were: COVID-19 autopsy reports; search results involving the selected terms COVID-19, Autopsy, Hyaluronan, Hyaluronic Acid, ARDS.

### Exclusion Criteria

Studies were excluded if they lacked (i) corresponding outcome parameters or research data or (ii) did not have available full text.

### Search Strategy

We conducted a targeted systematic review following the PRISMA flow diagram methodology. J.A.A. and M.A.M. systematically searched the electronic databases, PubMed and Scopus, for eligible reports following the PRISMA methodology (flow diagrams and search terms are listed in Figure 3). We included full reports with original data and applied no exclusions based on language. The search deadline was on May 27, 2020.

**Figure 1.**
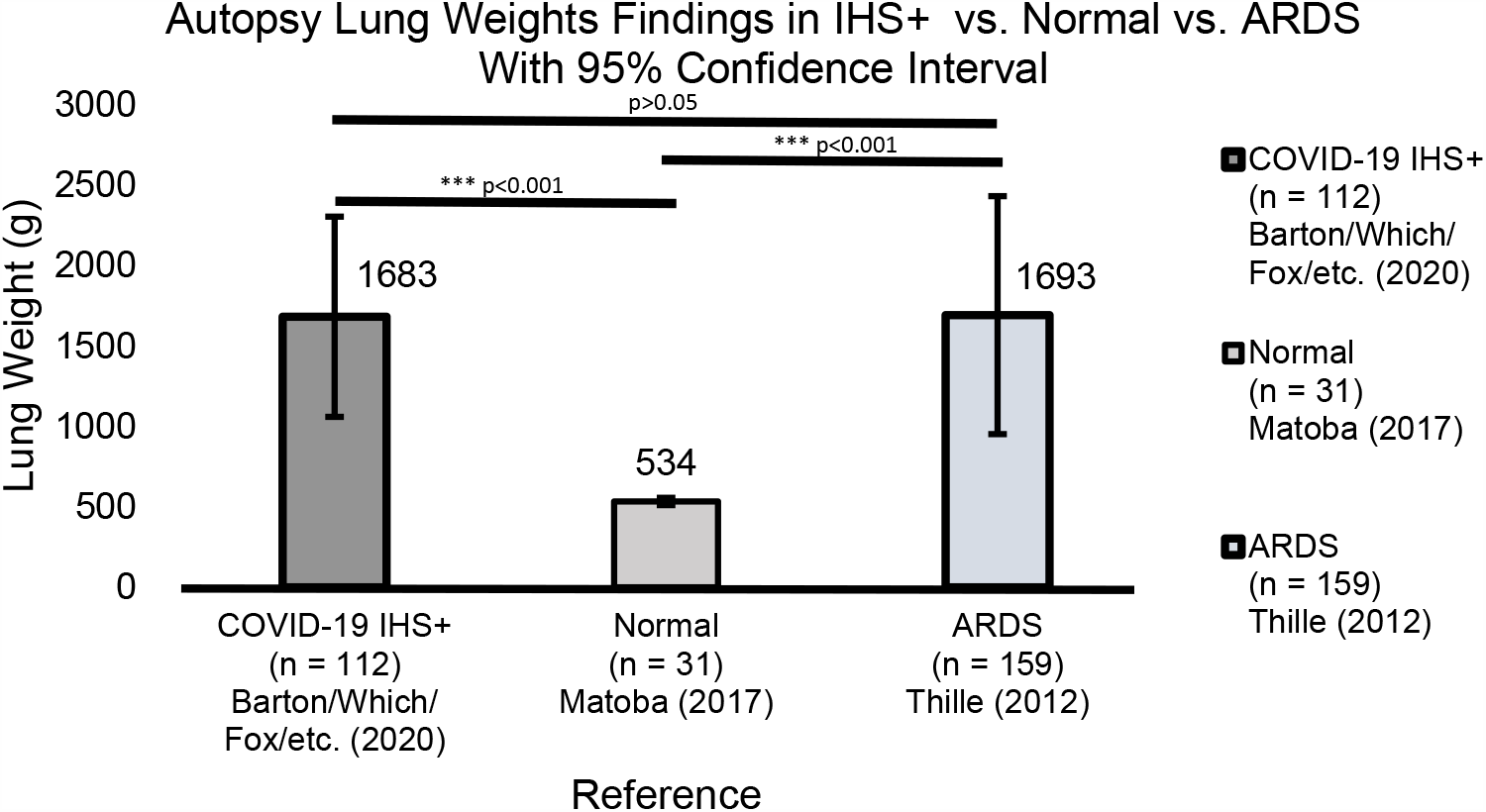
Statistical analysis of lung weights in IHS+, normal, and ARDS autopsies. A one-way ANOVA was performed followed by a post-hoc Tukey HSD.

**Figure 2.**
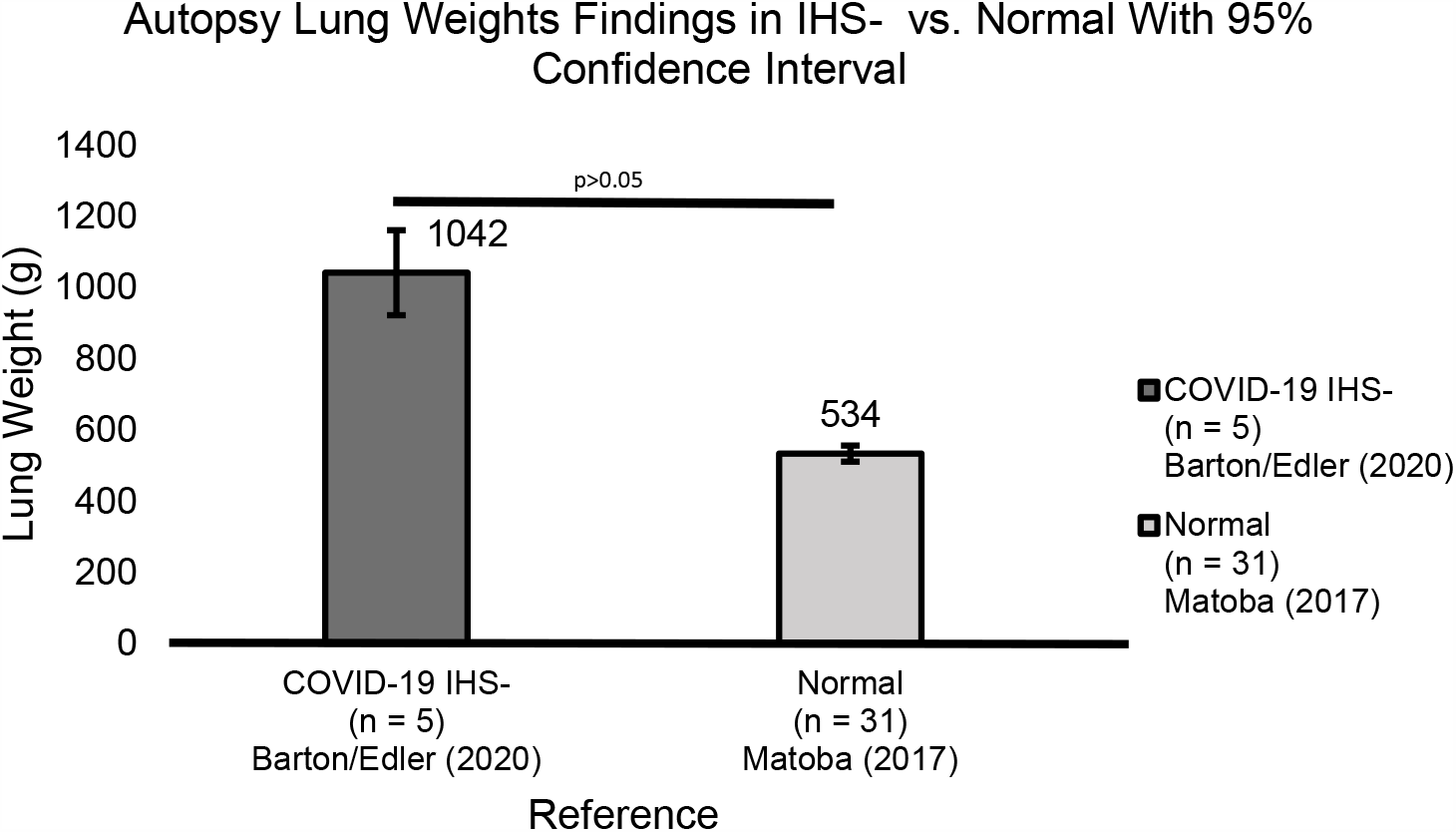
Statistical analysis of lung weights in IHS- and normal autopsies. A one-way ANOVA was performed followed by a post-hoc Tukey HSD. Because SARS-CoV-2 autopsy lung weights were limited, a small sample size was analyzed for IHS+ victims.

**Figure 3.**
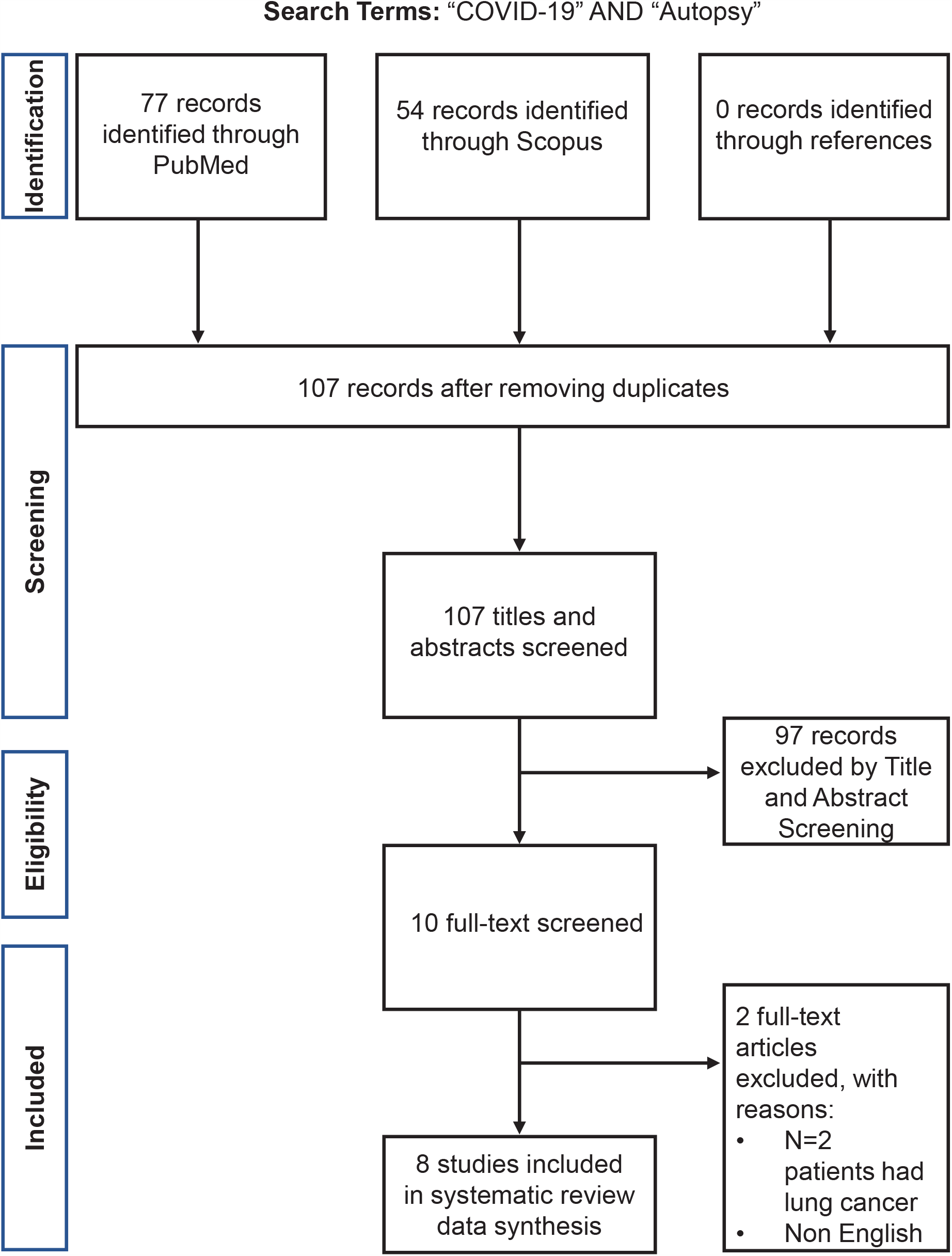
Flow chart of study selection

### Data Extraction

The elements extracted included sample size, location, measurement indicators of lung, spleen, liver, and circulatory systems. Literature was reviewed independently by two researchers (J.A.A. and M.A.M.). Autopsy reports were also screened for pre-existing conditions (n = 2 subjects with lung cancer and n = 1 non English literature cases were excluded). The primary outcome of interest was SARS-CoV-2 autopsy results with reference to lung weights. Mechanisms related to HA’s role in ARDS pathology were selected for detailed analysis.

### Data Collection and Quality Assessment

Relevant data elements were identified from each publication and recorded in Microsoft Excel (Microsoft Corporation, Redmond, WA, USA) and Zotero (George Mason University, Fairfax, VA, USA). All citations extracted from the PubMed or Scopus electronic databases were deemed to be of sufficient scientific quality.

### Data Synthesis and Analysis

We summarized the autopsy findings and compared the association between each outcome from the relevant studies. Further collating these results with citations related to HA and ARDS. These findings were compiled in an outcome status report in the form of a supplementary table, listing the significant findings from each case study. Statistical analysis was carried out with respect to both COVID-19, ARDS, and normal lung weights.

## Results

### Search Results

We identified 107 eligible publications after removing duplicates (Figure 3). Of these, 95 publications were excluded after screening titles and abstracts. Of the remaining, 12 publications were reviewed with full-text reads, two publications that did not meet the inclusion criteria were excluded, leaving 10 publications for the analysis. The final autopsy count as of May 27th, 2020 was 112 cases. All publications were considered sufficient scientific quality and the anatomic and histologic findings in SARS-CoV-2 reports were collated and further analyzed. Biological factors such as age ranged from 31 to 96, with an average age of 75. It was also found that 58% of the cases were male. Additionally, 51% of the victims had lung weights 3 times or greater than the normal value. Beyond, the average body mass index (BMI) was 28.5 kg/m^2^, we also found that 54% of patients were overweight or obese (BMI ≥ 25 kg/m^2^). Type II diabetes mellitus was present in 27% of cases and 40% of patients had a history of hypertension. We found that 36% of males and 26% of females were obese (BMI ≥ 30 kg/m^2^). Case 1, (Barton) with bilateral lung parenchymal swabs positive by rRT-PCR for SARS-CoV-2 had a lung weight of 2,452g - approximately 4.6 times the normal adult lung weight and showed the presence of hyaline membranes and bilateral diffuse alveolar damage.

Edler et al. identified 80 consecutive full autopsies that were segregated based on 4 categories (category 1: definite COVID-19 death; category 2: probable COVID-19 death; category 3: possible COVID-19 death with an equal alternative COD; category 4: COVID-19 detection with COD not associated to COVID-19)^[29]^. Autopsies which fell into category 4 were extracted and fed into the IHS-category (n=5). We found that the average lung weights of IHS-patients was 1042g with an average age of 71. Case 2, (Barton) autopsy revealed positive nasopharyngeal but negative bilateral lung parenchymal swabs by rRT-PCR for SARS-CoV-2, lung weight was documented at a normal 1191g^[9]^.

### Statistical Analysis

Average lung weights^[3, 9, 21-26]^ were plotted on MATLAB (MathWorks, Natick, MA, USA) and Microsoft Excel (Microsoft Corporation, Redmond, WA, USA). A one-way Analysis of Variance (ANOVA) coupled with a post-hoc analysis was performed using a Tukey Honestly Significant Difference (HSD) test. Namely, a Tukey HSD test was performed to adjust the p-values and indicate group statistical significance; however, note that a Tukey HSD test requires a studentized range distribution. This statistical model was performed using a 95% confidence interval between the groups (α = 0.05). There was a limited number of lung weights in confirmed COVID-19 IHS-patients, we found no statistical significance between IHS-patients and normal controls (Figure 2). One COVID-19 IHS-case was recorded with a combined lung weight of 1191g^[9]^.

## Discussion

We employed a systems biology^[8]^, rather than a reductionist approach, to better characterize and understand the mechanisms associated with the morbidity and mortality associated with COVID-19. We propose that a hyperacute rise in HA concentration is the key driver of morbidity and mortality in SARS-CoV-2 patients. The induced, sudden, and overwhelming increase in HA concentration, directly leads to a fatal accumulation of water in the lung parenchyma, overpowering the body’s defense mechanisms, causing rapid asphyxia and death, similar to drowning. No interventions directed towards the down stream effects of this massive accumulation of HA, and requisite water, can prevent the consequences of this singular hydrostatic event.

Under appreciated and neglected since the first description of the clinical features of ARDS based on careful autopsy results in 1967^[4]^, the morphologic and spacial implications of the sudden outpouring of HA and it’s sequestration of water in the alveolar environment, has not been fully appreciated and specifically addressed. Unfortunately, the historical focus has been centered on the down stream consequences, and not the critical and key physical dimensions and spacial aspects of a large volume of accumulated and misplaced water in the lung. Essentially the lung parenchyma, as a direct consequence of massive increases in HA, rapidly becomes saturated with water. Occurring within minutes, these shifts in water, as a direct result of a variety of injuries, and in this case, yet unknown and particularly lethal aspects of the SARS-CoV-2 infection, result in ARDS and require ventilation within hours.

Ding et al, performed a correlation analysis of the severity and clinical prognosis of 32 cases of patients with COVID-19 who were divided into three groups denoted as critical (n=11), severe (n=10), and mild (n=11) groups upon admission^[28]^. A comparison of fibrosis indicators (i.e. hyaluronic acid, type III/IV procollagen, and laminin) was performed among the three aforementioned groups. When analyzing their clinical findings of HA as a fibrosis indicator, we found that HA was significantly elevated in the critical group compared to the other two groups (Figure 7)^[28]^. We believe that this is a strong indication of the physico-chemical and biochemical role of HA in the progression of COVID-19. We maintain that HA is an important driver of ARDS and a potential prognostic bio marker that needs to be investigated further to understand the course of the morbidity and morality in COVID-19 victims.

**Figure 4.**
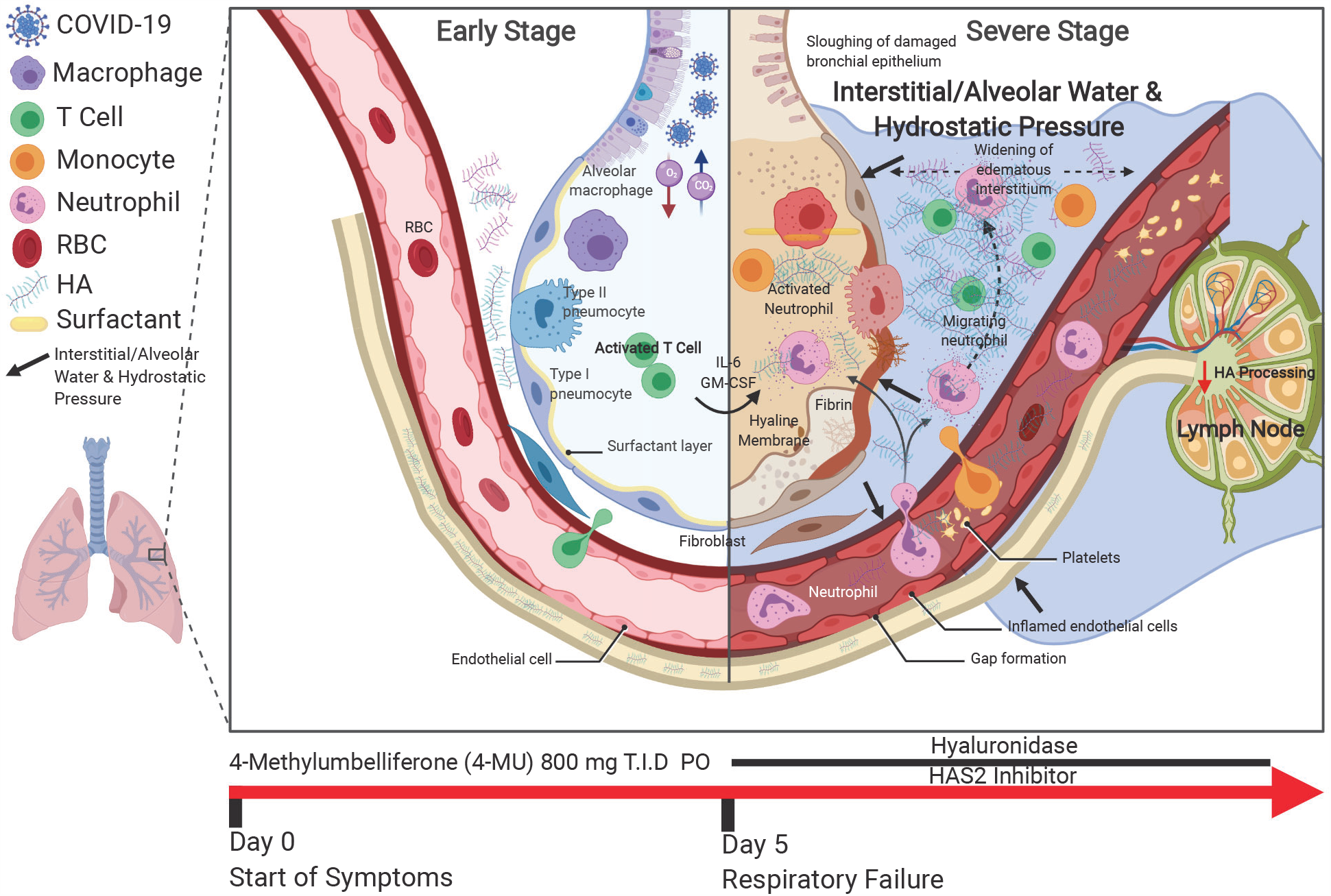
Schematic of the progression of SARS-CoV-2 infection and potential interventions. Inhibition of hyaluronan synthase and elimination of hyaluronan should be considered.

**Figure 5.**
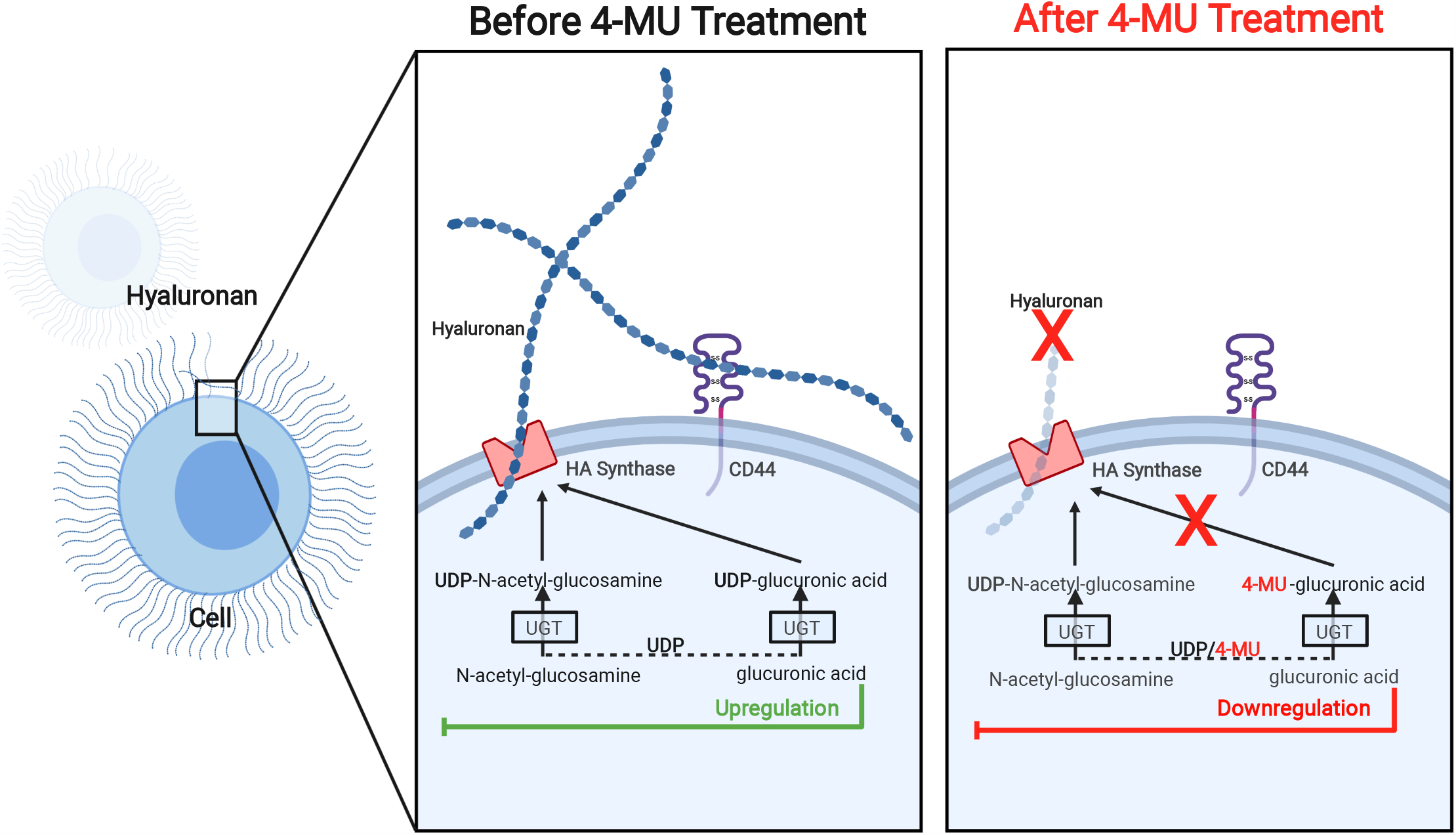
Schematic of the 4-MU treatment and biochemical pathway. Inhibition of hyaluronan synthase and elimination of hyaluronan should be considered.

**Figure 6.**
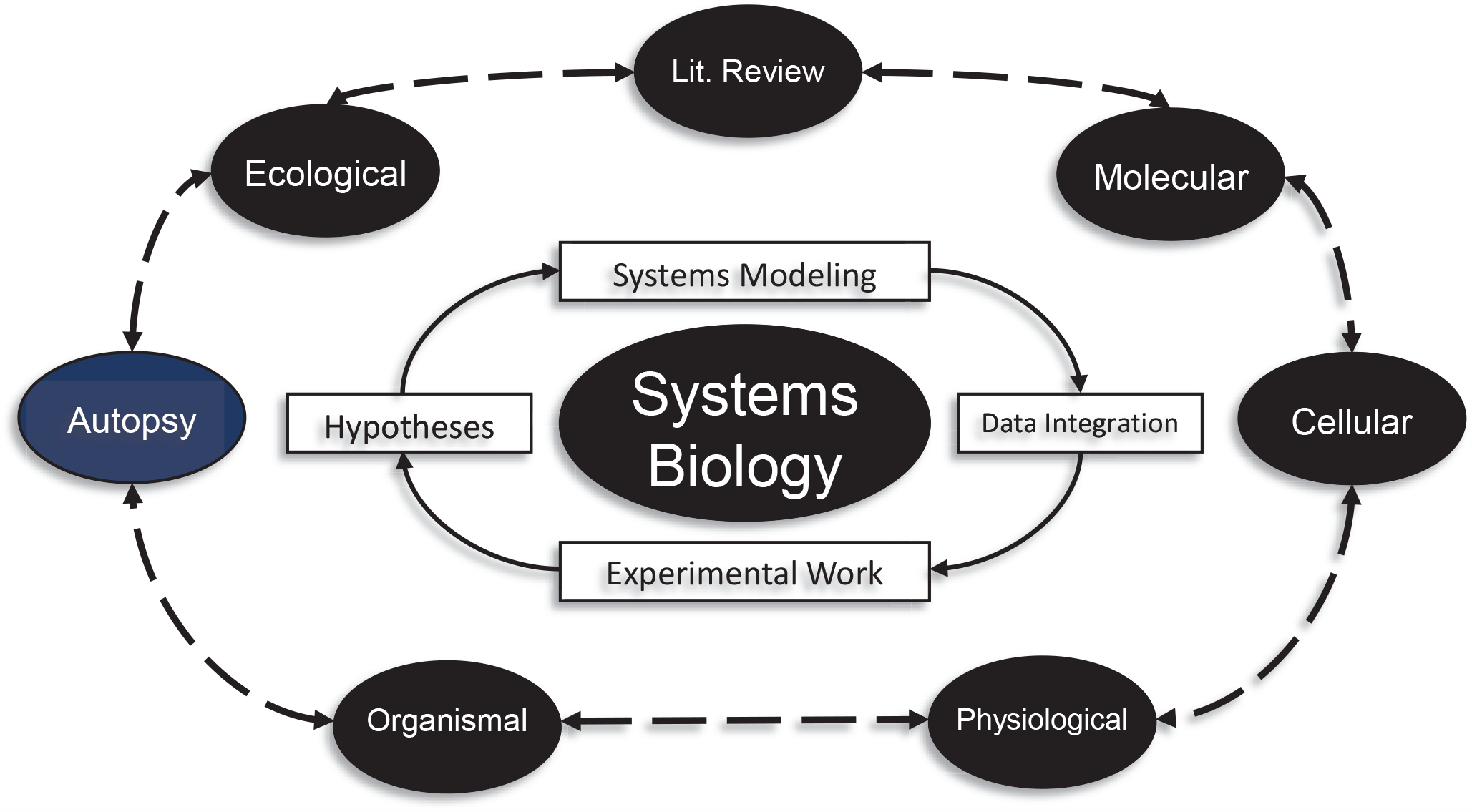
Theoretical framework outlining the elements of a systems biology, in contrast to reductionist, approach to complex disease states. Note the inclusion of autopsy as an integral component of this dynamic network.

**Figure 7.**
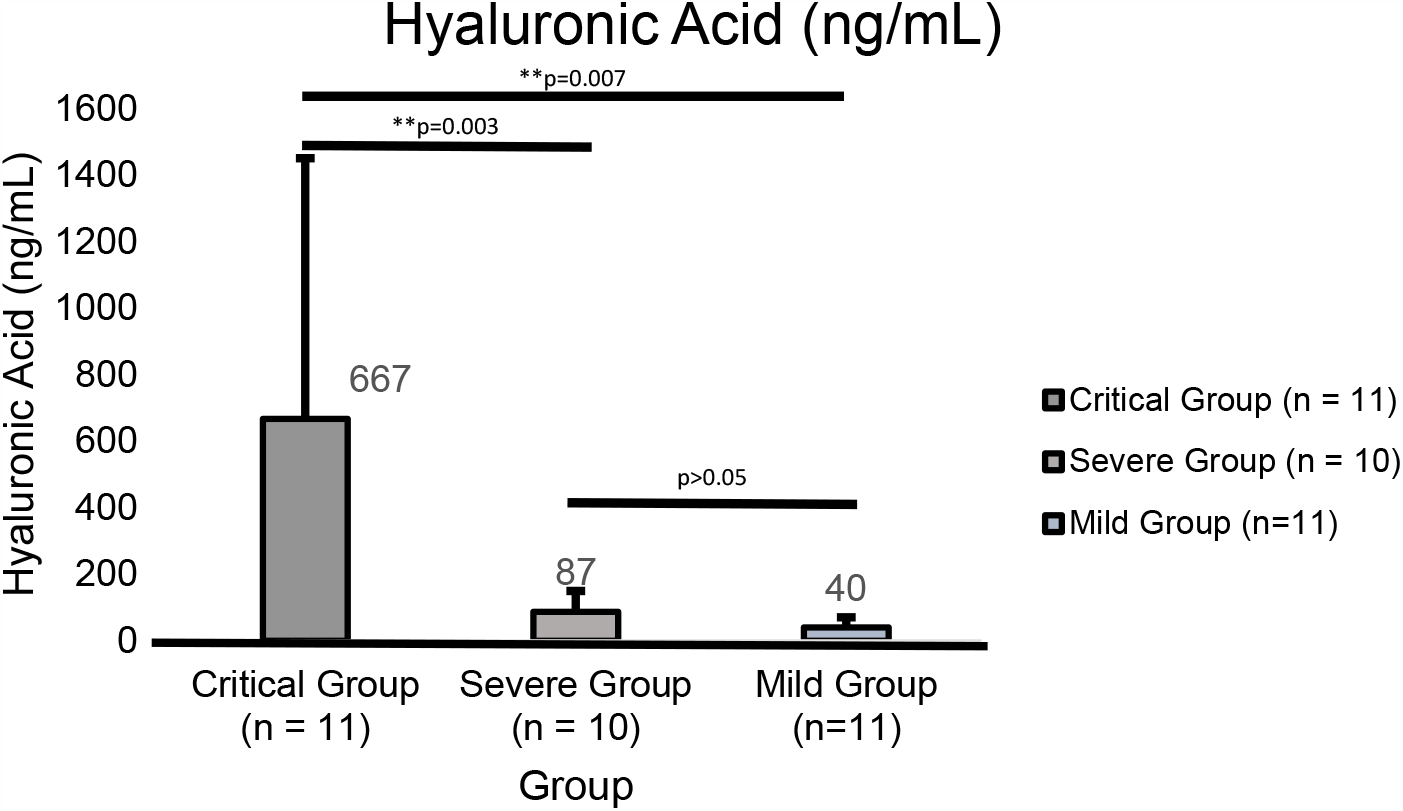
From the laboratory indicators of the three groups of COVID-19 patients at admission, hyaluronic acid was higher in the critical group than in the other two groups.

Critically, with respect to the above mechanism, it follows that conventional treatment with positive pressure ventilation, and anti-inflammatory medications can not overcome the physical effects of water saturated lung tissue driven by the rapid and early accumulation of HA in high-risk individuals.

These findings, and others, suggest that treatments targeting the production of HA may be life-saving in the present pandemic. We hypothesize that the most beneficial use of 4-MU in pre-symptomatic high risk patients at the time of initial diagnosis or exposure in high risk setting such as nursing homes. In addition the use of 4-MU might be considered in patients that are currently on ventilators, and certainly in symptomatic patients who have not crossed the line into respiratory failure, is strongly suggested by the extant literature.

Fortunately, evidence exists that 4-MU, which has been readily available in European Union as an over the counter product, called Cantabilin®, with a current marketing authorization via the Italian Medicines Agency (AIC no. 02130002) might be available for compassionate use.

Given the weight of these findings as well as the long track record of the safe use of 4-MU, and particularly the pressing need and current lack of a specific treatment for the COVID-19 ARDS phenotype, we propose that those on the front lines consider immediate use of 4-MU under compassionate use guidelines.

The work of Paul Bollyky, M.D., PhD. and Nadine Nagy PhD. et al., would suggest that 400-800 mg by mouth three times per day at first sign of infection in confirmed high risk patients might be reasonable initial doses^[18]^. These findings provide a rationale for studying the use of 4-MU in COVID-19 induced hyaluronan storm in a prospective, double-blinded clinical trial in high-risk patients. We strongly recommend these trials starting at the earliest time frame possible, and encourage readers to recommend specific protocols for doing so. It is known that drugs have unanticipated and unwanted side effects, therefore precautions must be taken in the clinical application of this, or any other compound for the treatment of COVID-19. Certainly, 4-MU should only be used under the direct supervision of skilled physicians.

## Conclusion

COVID-19 disease has resulted in a disastrous pandemic of enormous and compounding consequences, the likes of which have not been seen in modern times. We propose that the induced hyaluronan storm syndrome (IHS), is the model that best addresses the heretofore perplexing respiratory failure that is the proximal cause of death in a minority, but ever rising number, of patients. We encourage researchers and clinicians to put our model to the test, and expand upon our early understanding of this disease. In addition to treating and preventing IHS in currently infected individuals now; an aggressive research effort should be undertaken to discover why the majority of individuals who are exposed to the virus are either minimally or asymptomatic, while a minority of high-risk individuals rapidly progress to respiratory failure and death. The answer to this question will have profound implications for our fundamental understanding and approach to disease, and for the individuals and institutions charged with the management of this and future threats to global health and well-being. Foundations have been shaken, and the future will be profoundly shaped by these historic events, as history has been shaped by similar events in the past. Let us hope our collective responses are enlightened, cooperative, and well reasoned.^[20]^ Future directions include the reporting of further IHS± SARS-CoV-2 lung weights and performing additional autopsies on SARS-CoV-2 victims. We recommend the measurement of HA in serum as a potential indicator of IHS status.

## Data Availability

The data that support the findings of this study are openly available in publicly available electronic databases.
[1] Burton JL. A bite into the history of the autopsy. Forens Sci Med Pathol. 2005 Dec 1;1(4):277 84.
[2] Bassat Q, Castillo P, Alonso PL, Ordi J, Menendez C. Resuscitating the Dying Autopsy. PLoS Med [Internet]. 2016 Jan 12 [cited 2020 Apr 10];13(1). Available from: https://www.ncbi.nlm.nih.gov/pmc/articles/PMC4710495/
[3] Matoba K, Hyodoh H, Murakami M, Saito A, Matoba T, Ishida L, et al. Estimating normal lung weight measurement using postmortem CT in forensic cases. Leg Med (Tokyo). 2017 Nov;29:77:81.
[4] Ashbaugh DavidG, Boyd Bigelow D, Petty ThomasL, Levine BernardE. ACUTE RESPIRATORY DISTRESS IN ADULTS. The Lancet. 1967 Aug 12;290(7511):319:23.
[5] Bhatraju PK, Ghassemieh BJ, Nichols M, Kim R, Jerome KR, Nalla AK, et al. Covid 19 in Critically Ill Patients in the Seattle Region Case Series. New England Journal of Medicine. 2020 Mar 30;0(0)
[6] Konig MF, Powell M, Staedtke V, Bai R Y, Thomas DL, Fischer N, et al. Targeting the catecholamine cytokine axis to prevent SARS CoV 2 cytokine storm syndrome. medRxiv. 2020 Apr 8;2020.04.02.20051565.
[7] Huang C, Wang Y, Li X, Ren L, Zhao J, Hu Y, et al.Clinical features of patients infected with 2019 novel coronavirus in Wuhan, China.Lancet. 2020;395:497:506.
[8] Tavassoly I, Goldfarb J, Iyengar R. Systems biology primer: the basic methods and approaches. Kolch W, Fey D, Ryan CJ, editors. Essays in Biochemistry. 2018 Oct 26;62(4):487:500.
[9] Barton LM, Duval EJ, Stroberg E, Ghosh S, Mukhopadhyay S. COVID 19 Autopsies, Oklahoma, USA. Am J Clin Pathol [Internet]. [cited 2020 Apr 16]; Available from: https://academic.oup.com/ajcp/article/doi/10.1093/ajcp/aqaa062/5818922
[10] Modig J, Hallgren R. Increased hyaluronic acid production in lung a possible important factor in interstitial and alveolar edema during general anesthesia and in adult respiratory distress syndrome. Resuscitation. 1989 Jun;17(3):223:31.
[11] Lauer ME, Dweik RA, Garantziotis S, Aronica MA. The Rise and Fall of Hyaluronan in Respiratory Diseases [Internet]. Vol. 2015, International Journal of Cell Biology. Hindawi; 2015 [cited 2020 Apr 4]. Available from: https://www.hindawi.com/journals/ijcb/2015/712507/
[12] Johnson P, Arif AA, Lee Sayer SSM, Dong Y. Hyaluronan and Its Interactions With Immune Cells in the Healthy and Inflamed Lung. Front Immunol [Internet]. 2018 [cited 2020 Apr 17];9. Available from: https://www.frontiersin.org/articles/10.3389/fimmu.2018.02787/full
[13] Bell TJ, Brand OJ, Morgan DJ, Salek Ardakani S, Jagger C, Fujimori T, et al. Defective lung function following influenza virus is due to prolonged, reversible hyaluronan synthesis. Matrix Biol. 2019 Jul;80:14:28.
[14] McKallip RJ, Fisher M, Do Y, Szakal AK, Gunthert U, Nagarkatti PS, et al. Targeted Deletion of CD44v7 Exon Leads to Decreased Endothelial Cell Injury but Not Tumor Cell Killing Mediated by Interleukin 2 activated Cytolytic Lymphocytes. J Biol Chem. 2003 Oct 31;278(44):43818:30.
[15] McKallip RJ, Hagele HF, Uchakina ON. Treatment with the Hyaluronic Acid Synthesis Inhibitor 4 Methylumbelliferone Suppresses SEB Induced Lung Inflammation. Toxins (Basel). 2013;5(10):1814:1826. doi:10.3390/toxins5101814
[16] Bray BA, Sampson PM, Osman M, Giandomenico A, Turino GM. Early Changes in Lung Tissue Hyaluronan (Hyaluronic Acid) and Hyaluronidase in Bleomycin induced Alveolitis in Hamsters. American Review of Respiratory Disease [Internet]. 2012 Dec 17 [cited 2020 Apr 19]; Available from: https://www.atsjournals.org/doi/abs/10.1164/ajrccm/143.2.284
[17] Reeves SR, Barrow KA, Rich LM, White MP, Shubin NJ, Chan CK, et al. Respiratory Syncytial Virus Infection of Human Lung Fibroblasts Induces a Hyaluronan Enriched Extracellular Matrix That Binds Mast Cells and Enhances Expression of Mast Cell Proteases. Front Immunol [Internet]. 2020 Jan 28 [cited 2020 Mar 28];10. Available from: https://www.ncbi.nlm.nih.gov/pmc/articles/PMC6997473/
[18] Nagy N, Kuipers HF, Frymoyer AR, et al. 4 Methylumbelliferone Treatment and Hyaluronan Inhibition as a Therapeutic Strategy in Inflammation, Autoimmunity, and Cancer. Frontiers in Immunology. 2015;6. doi:10.3389/fimmu.2015.00123
[19] Liu Q, Wang RS, Qu GQ, Wang YY, Liu P, Zhu YZ, et al. Gross examination report of a COVID 19 death autopsy. Fa Yi Xue Za Zhi. 2020 Feb;36(1):21:3.
[20] Gully PR. Pandemics, regional outbreaks, and sudden onset disasters. Healthc Manage Forum. 2020 Feb 5;0840470420901532.
[21] Sekulic, M., Harper, H., Nezami, B. G., Shen, D. L., Sekulic, S. P., Koeth, A. T., Harding, C. V, Gilmore, H., & Sadri, N. (2020). Molecular Detection of SARS CoV 2 Infection in FFPE Samples and Histopathologic Findings in Fatal SARS CoV 2 Cases. American Journal of Clinical Pathology, 1:11. https://doi.org/10.1093/ajcp/aqaa091
[22] Aguiar, D., Lobrinus, J. A., Schibler, M., Fracasso, T., & Lardi, C. (2020). Inside the lungs of COVID 19 disease. International Journal of Legal Medicine. https://doi.org/10.1007/s00414 020 02318 9
[23] Fox, S. E., Akmatbekov, A., Harbert, J. L., Li, G., Brown, J. Q., & Heide, R. S. Vander. (2020). Articles Pulmonary and cardiac pathology in African American patients with COVID 19 : an autopsy series from New Orleans. The Lancet Respiratory, 2600(20), 1:6. https://doi.org/10.1016/S2213 2600(20)30243 5
[24] Ackermann, M., Verleden, S. E., Kuehnel, M., Haverich, A., Welte, T., Laenger, F., Vanstapel, A., Werlein, C., Stark, H., Tzankov, A., Li, W. W., Li, V. W., Mentzer, S. J., & Jonigk, D. (2020). Pulmonary Vascular Endothelialitis, Thrombosis, and Angiogenesis in Covid 19. New England Journal of Medicine, NEJMoa2015432. https://doi.org/10.1056/NEJMoa2015432
[25] Wichmann, D., Sperhake, J.P., Lutgehetmann, M., Steurer, S., Edler, C., Heinemann, A., Heinrich, F., Mushumba, H., Kniep, I., Schroder, A. S., Burdelski, C., de Heer, G., Nierhaus, A., Frings, D., Pfefferle, S., Becker, H., Bredereke Wiedling, H., de Weerth, A., Paschen, H.R., Kluge, S. (2020). Autopsy Findings and Venous Thromboembolism in Patients With COVID 19. Annals of Internal Medicine, 25(4). https://doi.org/10.7326/m20 2003
[26] Buja LM, Wolf DA, Zhao B, Akkanti B, McDonald M, Lelenwa L, et al. The emerging spectrum of cardiopulmonary pathology of the coronavirus disease 2019 (COVID-19): Report of 3 autopsies from Houston, Texas, and review of autopsy findings from other United States cities. Cardiovascular Pathology. 2020 Oct;48:107233.
[27] Michael A. Mong, Jacob A. Awkal, P. E. M. (2020). Accelerated hyaluronan concentration as the primary driver of morbidity and mortality in high-risk COVID-19 patients: with therapeutic introduction of an oral hyaluronan inhibitor in the prevention of Induced Hyaluronan Storm Syndrome

https://www.scopus.com

https://www.ncbi.nlm.nih.gov/pubmed/

## Abbreviations

COVID-19: novel coronavirus disease 2019
SARS-CoV-2: severe acute respiratory syndrome coronavirus 2
HA: hyaluronic acid or hyaluronan
IHS: induced hyaluronan storm
CSS: cytokine storm syndrome
SEB: staphylococcal enterotoxin B

## Acknowledgements

We thank the Lisa Barton, M.D., Ph.D., for personal communications reviewing specific COVID-19 autopsy findings. This manuscript has been released as a pre-print at Medrxiv, (Mong et al.).

---

**Table 1.** Summary of autopsy lung weights, demographic profiles, comorbodities, and clinical manifestations

| Date | Reference | Patient # | Age | Sex | Lung Weights | HT | DM | BMI | POD |
| --- | --- | --- | --- | --- | --- | --- | --- | --- | --- |
| 10-Apr | Barton | 1 | 77 | M | 2452 | Y | N | 31.8 | O |
| 6-May | Wichmann | 2 | 52 | M | 2075 | N | N | 38.8 | I |
|  |  | 3 | 70 | M | 2250 | N | N | 22.2 | I |
|  |  | 4 | 71 | M | 2725 | Y | N | 36.8 | I |
|  |  | 5 | 63 | M | 1480 | N | Y | 37.3 | I |
|  |  | 6 | 66 | M | 1800 | N | N | 25.3 | I |
|  |  | 7 | 54 | F | 550 | N | N | 29.6 | I |
|  |  | 8 | 75 | F | 1345 | N | N | 26.3 | I |
|  |  | 9 | 82 | M | 2100 | N | Y | 27.8 | I |
|  |  | 10 | 87 | F | 890 | N | N | 15.4 | I |
|  |  | 11 | 84 | M | 1360 | Y | Y | 20.7 | I |
|  |  | 12 | 85 | M | 3420 | Y | N | 30 | I |
|  |  | 13 | 76 | M | 2870 | N | N | 34.4 | I |
| 27-May | Fox | 14 | 44 | M | 2080 | Y | Y | 37.5 | I |
|  |  | 15 | 44 | F | 1550 | Y | Y | 47.76 | I |
|  |  | 16 | 63 | F | 1620 | Y | N | 56.97 | I |
|  |  | 17 | 76 | F | 1720 | Y | Y | 38.15 | I |
|  |  | 18 | 68 | M | 2090 | N | Y | 35.75 | I |
|  |  | 19 | 78 | F | 1330 | Y | N | 33.1 | I |
|  |  | 20 | 53 | F | 605 | Y | N | 28.5 | I |
|  |  | 21 | 60 | F | 1930 | Y | N | 44.8 | I |
|  |  | 22 | 66 | F | 1210 | Y | N | 28.3 | I |
|  |  | 23 | 78 | F | 1390 | N | Y | 34.3 | I |
| 21-May | Ackermann | 24 | 68 | F | 1660 | Y | N | 35 | I |
|  |  | 25 | 86 | M | 1740 | Y | Y | 26 | I |
|  |  | 26 | 96 | M | 1780 | Y | N | 23 | I |
|  |  | 27 | 78 | M | 1600 | Y | Y | 44 | I |
|  |  | 28 | 66 | M | 1650 | Y | N | 29 | I |
|  |  | 29 | 74 | M | 1720 | Y | Y | 27 | I |
|  |  | 30 | 81 | F | 1620 | Y | N | 26 | I |
| 28-Apr | Houston/Buja | 31 | 62 | M | 1590 | N | N | 33.8 | O |
|  |  | 32 | 34 | M | 1980 | Y | Y | 51.65 | O |
|  |  | 33 | 48 | M | 1980 | N | N | 35.2 | O |
| 26-May | Sekulic | 34 | 81 | M | 2240 | Y | Y | N/A | I |
|  |  | 35 | 54 | M | 2150 | Y | Y | 29.9 | I |
| 19-May | Aguir | 36 | 31 | F | 2220 | Y | Y | 61 | O |
| 15-May | Edler | 37 | 59 | M | N/A | N | N | N/A | I |
|  |  | 38 | 52 | M | 2075 | N | N | 38.8 | O |
|  |  | 39 | 70 | M | 2250 | N | N | 22.2 | I |
|  |  | 40 | 71 | M | 2725 | N | Y | 36.8 | I |
|  |  | 41 | 63 | M | 2470 | N | N | 37.3 | I |
|  |  | 42 | 66 | M | 1800 | N | N | 25.3 | I |
|  |  | 43 | 54 | F | 550 | N | Y | 29.6 | I |
|  |  | 44 | 75 | F | 1345 | N | N | 26.3 | I |
|  |  | 45 | 82 | M | 2100 | N | Y | 27.8 | I |
|  |  | 46 | 82 | F | 890 | N | N | 15.4 | I |

**Table 1** (Continued)
|  |  |  |  |  |  |  |  |  |  |
| --- | --- | --- | --- | --- | --- | --- | --- | --- | --- |
| 15-May | Edler | 47 | 84 | M | 1360 | Y | Y | 20.7 | I |
|  |  | 48 | 85 | M | 3420 | Y | N | 30 | I |
|  |  | 49 | 76 | M | 2870 | N | N | 34.4 | I |
|  |  | 50 | 70 | F | 1425 | N | Y | 41.7 | I |
|  |  | 51 | 90 | M | 800 | N | N | 20.9 | I |
|  |  | 52 | 93 | M | 1145 | Y | Y | 18.6 | I |
|  |  | 53 | 76 | M | 2390 | N | N | 27.7 | I |
|  |  | 54 | 85 | M | 1690 | N | N | 19.4 | I |
|  |  | 55 | 90 | M | 1260 | N | Y | 22.3 | O |
|  |  | 56 | 75 | F | 900 | Y | N | 24.8 | O |
|  |  | 57 | 77 | F | 1035 | N | N | 20.4 | O |
|  |  | 58 | 86 | M | 1900 | N | N | 22.3 | O |
|  |  | 60 | 87 | F | 970 | N | N | N/A | O |
|  |  | 61 | 85 | F | 1240 | N | N | 17.3 | O |
|  |  | 62 | 86 | F | 1280 | N | N | 18.3 | O |
|  |  | 63 | 93 | M | 1530 | N | Y | 19.4 | I |
|  |  | 64 | 77 | M | 2505 | N | N | 19.4 | I |
|  |  | 65 | 85 | F | 1050 | N | N | 18.1 | I |
|  |  | 66 | 88 | M | 2110 | N | N | 23.4 | I |
|  |  | 67 | 90 | M | 1890 | N | N | 26.9 | I |
|  |  | 69 | 70 | M | 2300 | Y | N | 19.8 | I |
|  |  | 70 | 76 | W | 1540 | Y | N | 21.7 | I |
|  |  | 71 | 51 | M | 2185 | N | N | 20.7 | O |
|  |  | 72 | 84 | M | 1115 | N | N | 24.4 | I |
|  |  | 73 | 57 | M | 2200 | N | N | 33.3 | I |
|  |  | 74 | 75 | M | 1020 | Y | N | 34.6 | I |
|  |  | 76 | 88 | F | 990 | N | N | 18.5 | N/A |
|  |  | 77 | 56 | M | 1220 | N | Y | 37 | I |
|  |  | 78 | 91 | F | 1010 | N | N | 20.5 | O |
|  |  | 79 | 90 | F | 760 | N | N | 17.2 | O |
|  |  | 80 | 79 | F | 1880 | N | N | 18.7 | I |
|  |  | 81 | 87 | F | 1015 | N | N | 16.4 | I |
|  |  | 82 | 86 | M | 1190 | N | Y | 25.5 | O |
|  |  | 83 | 72 | M | 3110 | N | N | 47.1 | I |
|  |  | 85 | 82 | F | 925 | N | N | 26.8 | I |
|  |  | 86 | 54 | F | 2610 | N | N | 34.6 | I |
|  |  | 87 | 93 | F | 1110 | Y | N | 17.6 | O |
|  |  | 88 | 80 | M | 1580 | N | N | 40.6 | O |
|  |  | 89 | 82 | F | 1230 | N | N | 22.8 | I |
|  |  | 90 | 89 | F | 1650 | N | N | 25.9 | I |
|  |  | 91 | 78 | F | 1600 | N | N | 45.6 | I |
|  |  | 92 | 78 | F | 1080 | N | N | N/A | I |
|  |  | 93 | 92 | M | 1490 | Y | N | 25.5 | O |
|  |  | 94 | 71 | M | 2315 | Y | N | N/A | I |
|  |  | 95 | 85 | M | 1380 | N | N | 23.2 | I |
|  |  | 96 | 88 | M | 2005 | N | N | 23.5 | I |
|  |  | 97 | 86 | F | 1420 | N | N | 21.3 | N/A |

Abbreviations HT; Hypertension, DM2; Type II Diabetes Mellitus, BMI; Body Mass Index, POD; Place of Death, A; In Hospital, O; Out Hospital

**Table 2.** Summary of autopsy lung weights, demographic profiles, comorbodities, and clinical manifestations in IHS-patients.

| Date | Reference | Patient # | Age | Sex | Lung Weights | HT | DM | BMI | POD |
| --- | --- | --- | --- | --- | --- | --- | --- | --- | --- |
| 15-May | Edler | 1 | 58 | M | 1050 | N | N | N/A | O |
|  |  | 2 | 73 | M | 1030 | N | N | 29.3 | O |
|  |  | 3 | 86 | M | 860 | Y | Y | N/A | O |
|  |  | 4 | 94 | F | 1080 | N | N | 21.4 | I |
| 10-Apr | Barton | 5 | 42 | M | 1191 | N | N | 31.3 | I |

Abbreviations HT; Hypertension, DM2; Type II Diabetes Mellitus, BMI; Body Mass Index, POD; Place of Death, A; In Hospital, O; Out Hospital

## Notes

### Competing Interest Statement

In 2020, Michael Mong filed patent application number 63005439-Systems And Methods For Treating Corona Virus.

### Funding Statement

No external funding was acquired for this manuscript.

